# Machine learning based prognostic model and mobile application software platform for predicting infection susceptibility of COVID-19 using health care data

**DOI:** 10.1101/2020.10.09.20165431

**Authors:** R Srivatsan, Prithviraj N Indi, Swapnil Agrahari, Siddharth Menon, S Denis Ashok

**Affiliations:** VIT University, Vellore, Tamil Nadu, 632014, India

**Keywords:** Machine Learning, Prognostics, COVID-19, infection susceptibility, mobile application, random forests, support vector regression

## Abstract

From public health perspectives of COVID-19 pandemic, accurate estimates of infection severity of individuals are extremely valuable for the informed decision making and targeted response to an emerging pandemic. This paper presents machine learning based prognostic model for providing early warning to the individuals for COVID-19 infection using the health care data set. In the present work, a prognostic model using Random Forest classifier and support vector regression is developed for predicting the Infection Susceptibility Probability (ISP) score of COVID-19 and it is applied on an open health care data set containing 27 field values. The typical fields of the health care data set include basic personal details such as age, gender, number of children in the household, marital status along with medical data like Coma score, Pulmonary score, Blood Glucose level, HDL cholesterol etc. An effective preprocessing method is carried out for handling the numerical, categorical values (non-numerical), missing data in the health care data set. The correlation between the variables in the health care data is analyzed using the correlation coefficient and heat map with a color code is used to identify the influencing factors on the Infection Susceptibility Probability (ISP) score of COVID-19. Based on the accuracy, Precision, Sensitivity and F-scores, it is noted that the random forest classifier provides an improved classification performance as compared to Support vector regression for the given health care data set. Android based mobile application software platform is developed using the proposed prognostic approach for enabling the healthy individuals to predict the susceptibility infection score of COVID-19 to take the precautionary measures. Based on the results of the proposed method, clinicians and government officials can focus on the highly susceptible people for limiting the pandemic spread

**Methods:** In the present work, Random Forest classifier and support vector regression techniques are applied to a medical health care dataset containing 27 variables for predicting the susceptibility score of an individual towards COVID-19 infection and the accuracy of prediction is compared. An effective preprocessing is carried for handling the missing data in the health care data set. Correlation analysis using heat map is carried on the health care data for analyzing the influencing factors of Infection Susceptibility Probability (ISP) score of COVID-19. A confusion matrix is calculated for understanding the performance of classification of the based on the number of True-Positives, True-Negatives, False-Positives and False-Negatives. These values further used to calculate the accuracy, Precision, Sensitivity and F-scores.

**Results:** From the classification results, it is noted that the Random Forest classifier provides an classification accuracy of 99.7% precision of 99.8%, sensitivity of 98.8% and F-score of 99.29% for the given medical data set.

**Conclusion:** Proposed machine learning approach can help the individuals to take additional precautions for protecting people from the COVID-19 infection, clinicians and government officials can focus on the highly susceptible people for limiting the pandemic spread.

**Abbreviation Table:** 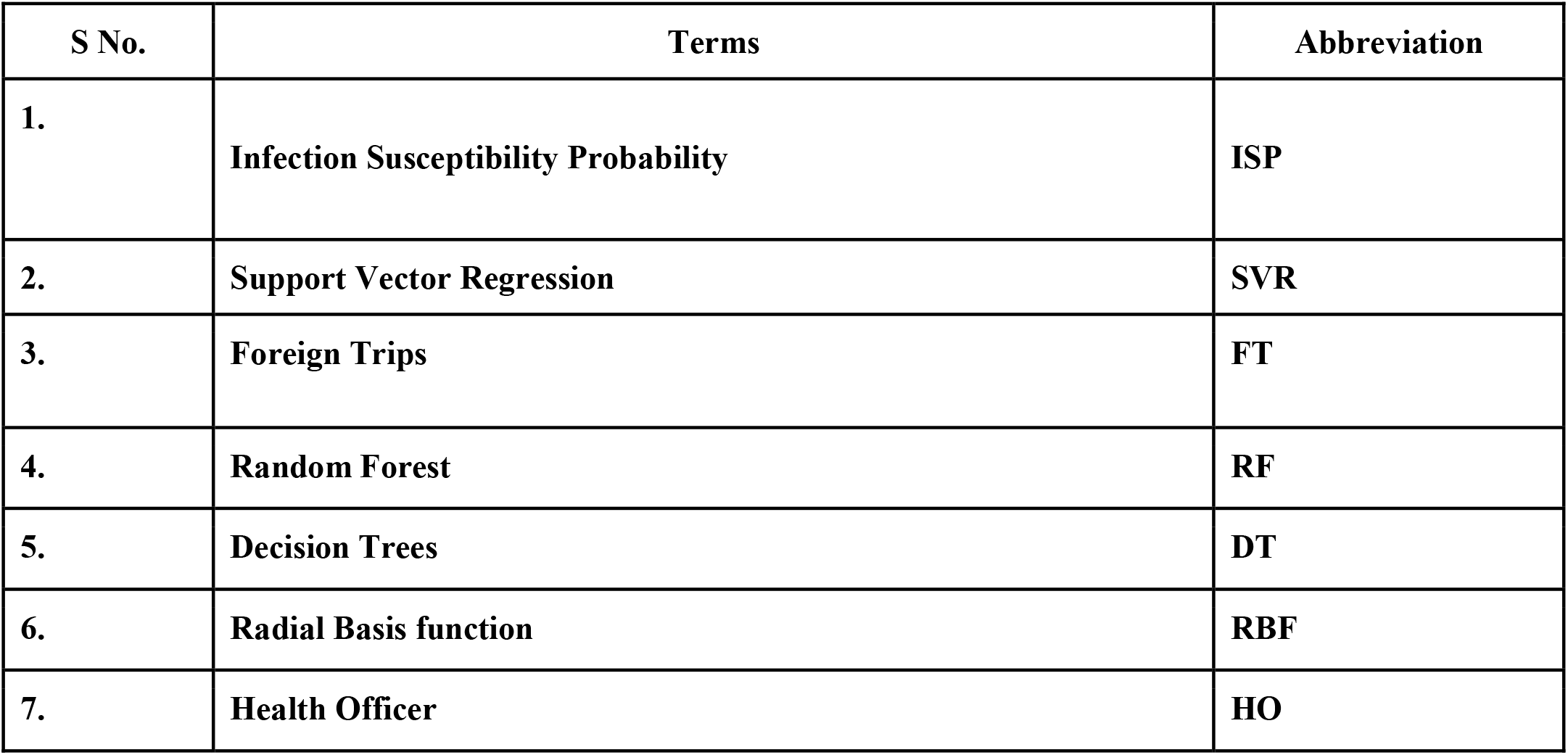

## Introduction

The recent outbreak of coronavirus disease 2019 (COVID-19) has created a great challenge for the healthcare system (Hui etal, 2020). Considering the lethal nature of COVID-19 outbreak and its worldwide spread, World Health Organization (WHO) and Centers for Disease Control and Prevention (CDC) at different nations have provided provisional guidelines for protecting people from getting affected and preventing the further spread of COVID-19 virus from infected individuals. RT-PCR tests from deep nasotracheal samples and Chest CT scan are commonly used for definitive diagnosis of COVID-19. (Repici A et al. 2020). Due to the quick spread of COVID-19, physicians in the health care systems are facing extreme difficulty in the physical examination and analysis of subsequent para clinical health care data for the accurate diagnosis of COVID-19. Hence, it is necessary develop software tools for easier way for interpreting the large scale health data set which can help the government and healthcare officials for quicker decision making during the Covid-19 pandemic situations.

With the capability of interpreting the hidden and complex patterns from huge, noisy or complex data, Artificial intelligence and machine learning techniques can play a major role in combating the COVID-19 pandemics. Few works have been reported by the researchers on use of machine learning techniques for the prediction and diagnosis of epidemics (Wynants L et al, 2020). An artificial intelligence based rapid diagnosis approach for COVID-19 patients developed using the analysis of Chest X ray images (Mei et al, 2020). An artificial intelligence based prediction model of the epidemics trend of COVID-19 is proposed by Yang etal, 2020. Linear Regression model is used for time series prediction of COVID-19 outbreak (Pandey et al, 2020). Mechanistic models have been reported to predict COVID-19 outbreak in real time (Liu etal, 2020). K means algorithm is applied to categorize the countries based on the number of confirmed COVID-19 cases (Carrillo-Larco et al, 2020). XGBoost machine learning model is proposed to estimate the survival ratio of severely ill Covid-19 patients (Yan et al. 2020). A classification using Fourier and Gabor methods is applied on dataset of COVID-19 (Al-Karawi et al 2020). Multi Layered Perception (MLP), Adaptive Network-based Fuzzy Inference System is used for predicting (Metsky et al. 2020). Support vector machine is applied to detect severely ill COVID patients from mild symptom COVID patients (Tang et al.2020). Convolutional neural network frameworks have been proposed to detect COVID-19 from chest X-ray images. (Narin et al, 2020). A prediction model for the propagation analysis of the COVID-19 is proposed by Li etal 2020. An interpretable mortality prediction model for COVID-19 patients is developed using the health care data set (Lan etal, 2020).

It is found that many machine learning approaches has been successfully implemented for the prediction and diagnostic purposes of COVID-19 using the clinical and health care data. However, prognostic frame works for early prediction of COVID-19 infection are found to be limited which can be helpful to take proactive measures to combat the virus spread. Random forest and Support vector machine algorithms are found to be popular in achieving the satisfactory results for the different prediction applications. Hence, this paper presents Random forest and Support vector machine algorithms based prognostic approach for predicting the infection susceptibility score for each individual using the health care data. The novelty of the proposed approach is the identification of the infection susceptibility prior to infection so that the regulative and preventive rules can be made for the individuals.

## Methods

Recently, machine learning techniques have applied for prognostic applications like prediction of disease symptoms, risks, survivability, and recurrence [Adnan Qayyum et al, 2020]. In the present work, a machine learning based prognostic approach is presented for predicting the Infection Susceptibility Probability (ISP) score of COVID-19 and categorizing the healthy individuals as low, medium, high using open health care data set. Among the various machine learning techniques, random forest and support vector regression has been considered for the present application due to their superior classification performance than other techniques such as linear regression, neural networks. Proposed prognostic approach is further developed as a strategic decision making tool by involving a versatile data base and a mobile application which enables the healthy individuals for taking precautionary measures and also government officials in prioritizing the resources in the hospital settings. Fig. 1 shows the major elements of the proposed approach and it is described below:

- A health care survey data set includes the demographic, epidemiological characteristics, and underlying comorbidities of the individuals with the target classes of risk factors namely - High risk (66%-100%), Medium risk (33%-66%) and Low risk (0%-33%). As the collected health care data may contain the missing values during the data collection, an effective preprocessing is essential and it is carried out before it is applied to the machine learning models for the classification applications.
- Supervised machine learning techniques consisting of random forest, support vector regression, linear regression, and neural networks for predicting Infection Susceptibility Probability (ISP) score of COVID-19 using the labeled heath care data
- A centralized data collection system and android mobile application which can be useful for sharing the multi modal health care data and predicting the infection susceptibility score to the individuals, health care professionals, government administrative officials.

**Fig 1.**
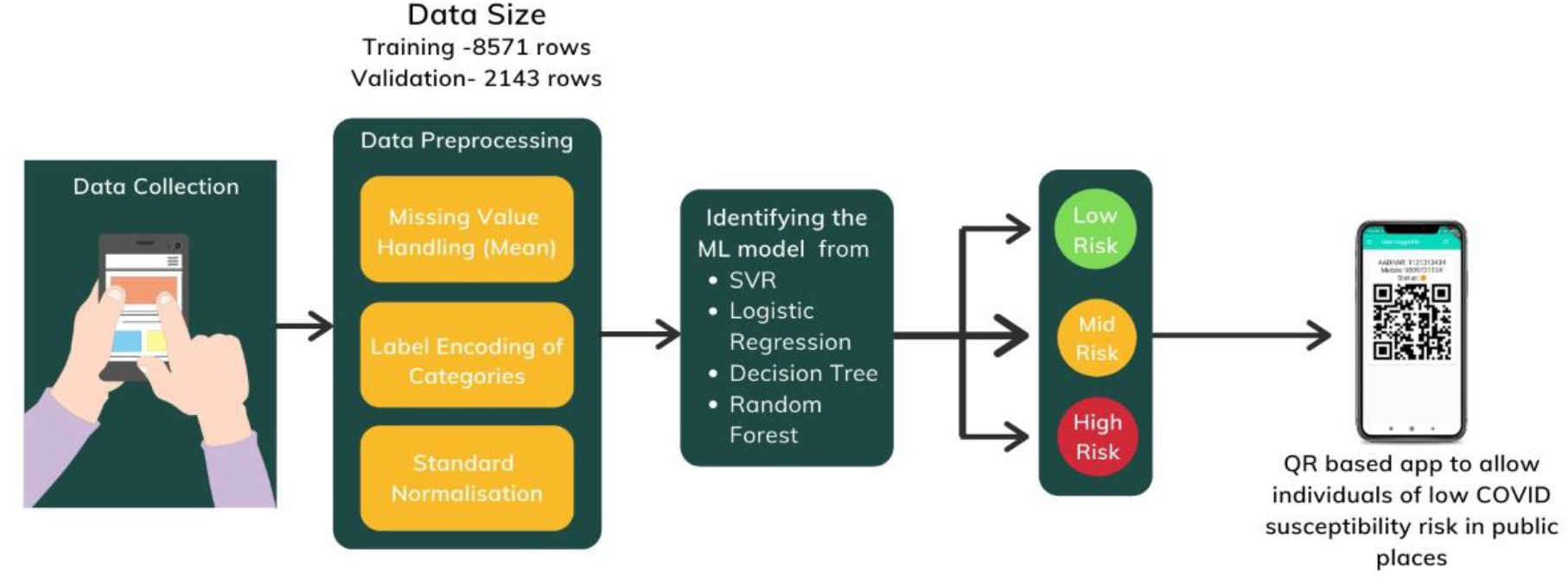
Proposed prognostic approach for predicting the susceptibility score of individuals using health care data

### Description of health care dataset

In the present work, open health care data set containing the demographic, epidemiological characteristics, and underlying comorbidities of the individuals is used for demonstrating the proposed prognostic approach and it available in the online repository Kaggle [Srijan,2020]. The data set contains 14498 rows and 27 columns. Table.1 shows the typical fields of the health care data set which include basic personal details such as age, gender, number of children in the household and marital status along with medical data like Coma score, Pulmonary score, Blood Glucose level, HDL cholesterol. Medical data chiefly includes comorbidity conditions such as Severe Acute Respiratory Infections (SARI), diabetes and heart syndromes. Vitals such as heart rate have also been considered in the modeling of the predictor infection susceptibility score of COVID-19.

**Table 1.** The various fields of the medical and health care data (Srijan Singh 2020)

| Variables | Description |
| --- | --- |
| people_ID | Unique ID for each person |
| Region | The area that the person belongs to |
| Gender | Gender of the person |
| Designation | Designation of the person |
| First_Name | Name of individual |
| Married | Marital status of individual |
| Children | Number of children in the family |
| Occupation | Sector of individual occupation |
| Mode_transport | Mode of transport that the individual frequently chooses to travel |
| cases/1M | Number of confirmed cases per 1 million population in that region |
| Deaths/1M | Number of Death case per 1 million population in that region |
| comorbidity | Co-occurring medical condition |
| Age | Age of the person |
| Coma score | Neurological coma score |
| Pulmonary score | Pulmonary PaO<sub>2</sub> (mmHg)/FiO<sub>2</sub> |
| cardiological pressure | Cardiological Mean systolic Arterial pressure (mmHg) |
| Diuresis | Diuresis in mL/Day |
| Platelets | Hematological Platelets 10/L |
| HBB | Hepatic Blood bilirubin (μmol/L) |
| d-dimer | d-dimer concentration in the blood (ng/ml) |
| Heart rate | number of times a person's heart beats per minute |
| HDL cholesterol | High-density lipoprotein level (milligrams per decilitre) |
| Charlson Index | index for a patient who may have any of the listed comorbid disease conditions |
| Blood Glucose | strength of glucose present in the blood (millimoles per litre) |
| Insurance | Medical Insurance spending cover (in Rs.) |
| salary | Annual salary of the individual |
| FT/month | Average foreign trips taken by the individual per month, considering last 2 year data |

### Data Preprocessing and preparation

It is noted that the health care survey data is multi modal as it contains many numerical, categorical values (non-numerical) and many machine learning algorithms cannot handle data in this form. Also, there can be missing values in the relevant fields of the data set which will reduce the accuracy of machine learning algorithms. In order to overcome these difficulties, data preprocessing and preparation is essential and it is carried out on the health care data set using Label encoding to achieve accurate results using the machine algorithms.

### Data Normalization

Standardization of the dataset makes a very crucial role in the pipeline of the ML model since if the individual features do not reassemble standard normal distribution of data points, the model would become erratic in its predictions. This involves a technique of reducing mean value from each individual data point and performing a scaling operation to them in order to obtain unit variance per cell of the data.

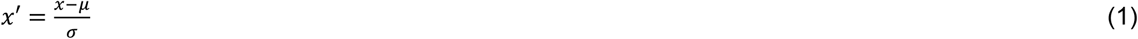

Where μ denotes data’s mean value and *σ* denotes the standard deviation obtained as square root of variance.

### Correlation analysis of health care data using Heat map

With many fields involving demographic, epidemiological characteristics, and underlying comorbidities of the individuals, a typical health care data set formulates a high dimensional feature space which has the strong and weak relevance to the Infection Susceptibility Probability (ISP) score. As the highly correlated features add noise and inaccuracy to the machine learning model, it is necessary to analyze the correlation between the variables in the health care data. In this work, correlation analysis of the health care data is carried using the correlation coefficient and heat map with a color code is used to visualize it. Based on the statistical analysis of health care data, correlation coefficient is calculated as the ratio of covariance and the standard deviation between two feature sets *a*_*i*_ and *b*_*i*_ is given by the following formula:

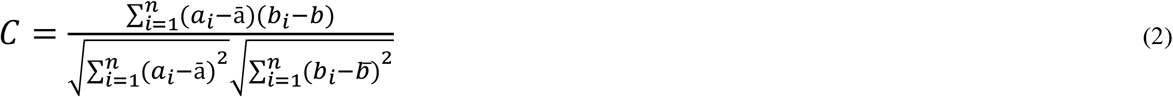

where *n* is the sample size, *a*_*i*_ and *b*_*i*_ are the *i*_*th*_ data values, and *a, b* are the mean values. The value of the coefficient (*C*) ranges between −1 and +1. The values of the correlation close to +1 indicate the strong positive correlation, those close to −1 show strong negative correlation, and those closest to 0 show no relation.

### Machine learning techniques for predicting the Infection Susceptibility Probability (ISP) score of COVID-19

Based on the statistical analysis of the health care data with the relevant information on the severity of the covid-19 infection, binary logistic regression model is fitted to the training sample and the coefficients of the regression model are used for calculating the probability of infection (Menelaos Pavlou et al, 2015), (Xinghuan Wang et al, 2020). In present work, supervised machine learning techniques such as random forest, support vector regression, linear regression, and neural networks have been applied for predicting and categorizing the susceptibility score of COVID-19 infection using the health care data. With the improved accuracy of classification, random forest and support vector regression is considered in the present work and it is explained below:

### Random Forest Classifier

With interpretable decision logic, Random forest algorithm (RF) is found to be one of the most promising classifier which uses multiple decision trees (DT) to train and predict data samples. The general structure of random forest with multiple decision trees is shown in Fig.2. The multiple ensemble DTs give rise to different classifications of infection susceptibility score. Here the value of *n* is chosen to be between 10 and 20 for optimum prediction. The majority scheme of vote is the terminal deciding factor of the model decision and throws the actual predicted class of the ISP of the individual as Low, medium, high.

**Fig 2.**
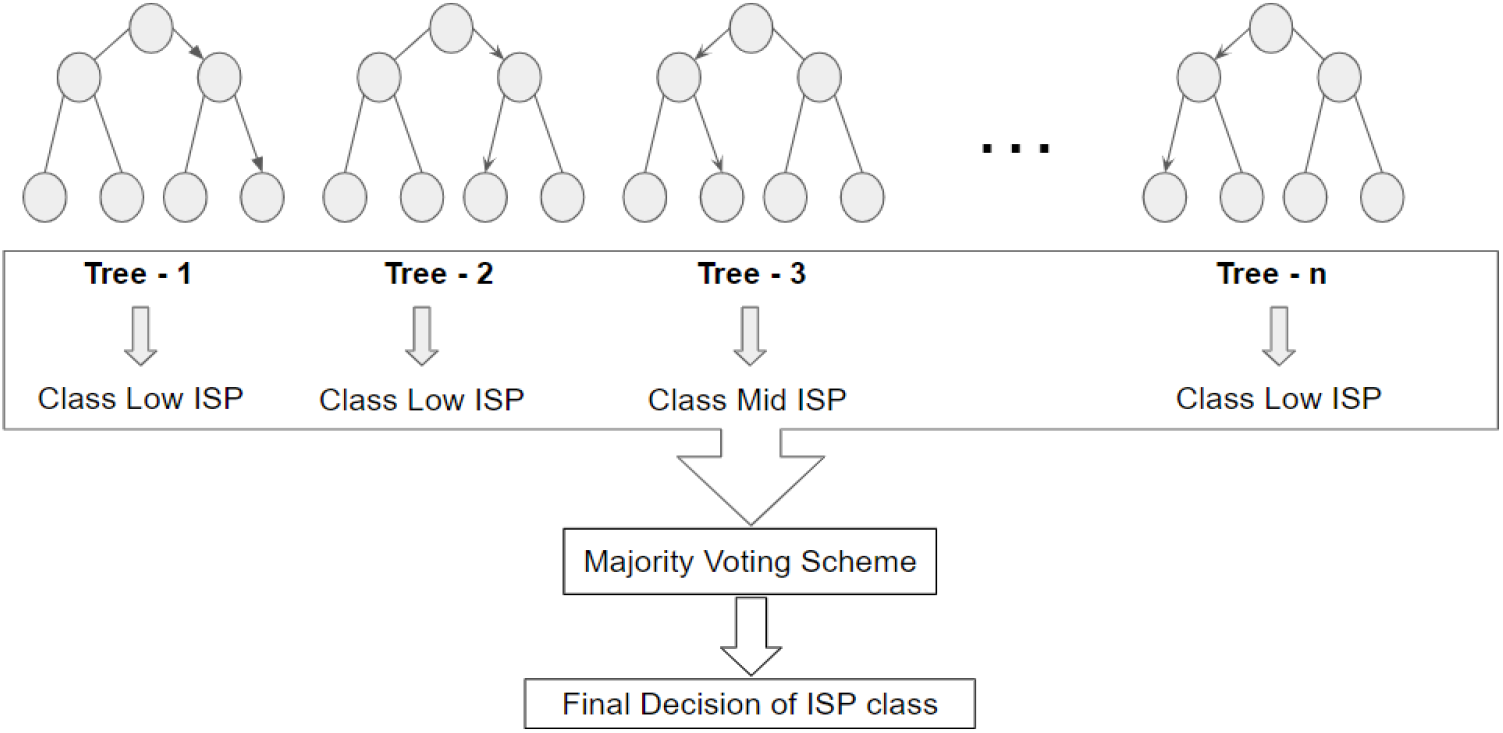
Illustration of the random forest architecture

In the random forests classification approach, the ensemble of Decision Trees (DT) is involved in calculating the GINI score as in equation (3).

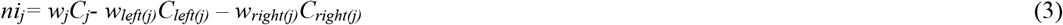

Here, the notations of the parameters are so following: ni_j_ refers to the significance of the node indexed j, W_j_ is indicating the weighted number of samples approaching the node indexed j, C_j_ indicates the impurity value of node indexed j, left(j) denotes the left child node from node indexed j and right(j) shows the right child node from node indexed j.

The second step is to obtain the importance given by each feature of the DT. This significance parameter can be computed using equation (4).

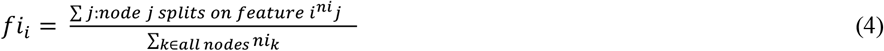

where fi_i_ denotes the importance of feature indexed i and the ni_j_ refers to the importance of node indexed j.

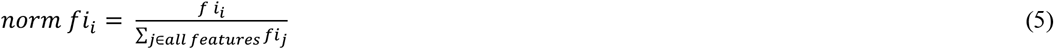

These features fi_i_ are now normalized using the equation

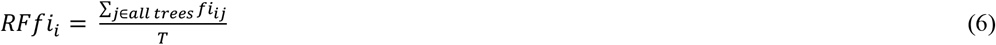

Then we can obtain the final feature of importance as mean of those of all the DTs as given in Fig.2, where RF f_i_ refers to the importance of feature indexed i computed through all DTs in the RF and norm fi_ij_ refers to the normalized feature significance parameter for index i in the DT indexed j and T indicates the total number of DTs.

### Support Vector Regression (SVR)

In the present work, support vector regression is applied for predicting the susceptibility score of COVID-19 infection as a continuous variable. Due to very high non linearity in the PCs, a Support Vector Regression based approach was chosen to be employed for obtaining probabilities of risk of infection using the medical data set.

Assuming that the set of training medical data *x*_*n*_ is a multivariate set of *N* observations with observed response values *y*_*n*_, a linear function is established as given below:

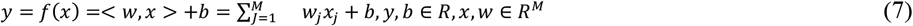

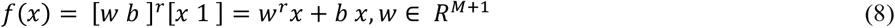

In the above equation x is a multidimensional input vector, with bias b and normal vector w. To ensure that it is as flat as possible, *f*(*x*) with the minimal norm value, a convex optimization problem is formulated to minimize the following:

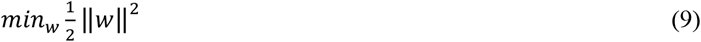

This shows that the normal vector should be approximated during the process. Magnitude of weights is usually interpreted as flatness to the function obtained in the computation.

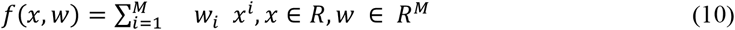

To minimize the loss between the actual and predicted value which is a major constraint SVR adopts epsilon-insensitive loss function. Although asymmetrical loss functions should be used to avoid underestimation and overestimation, the functions used are usually convex in nature.

Since most of COVID data is asymmetrical, linear methods wouldn’t provide accurate results. Nonlinear methods in Support Vector Regression (SVR) are handled by mapping the features to higher dimensional space called kernels.

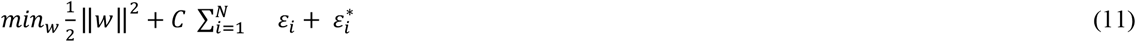

To achieve higher accuracy we replace all instances of x with K (xi,xj) from the earlier linear formula which leads to primal formulation shown in the above equation. The transformation of features to kernel space is shown in the above equation.

### Prototype Implementation of the Proposed Approach using a Mobile Application

In order to demonstrate the proposed approach for predicting the Infection Susceptibility Probability (ISP) score of COVID-19 of healthy individual, a mobile application software tool with user interface is developed for collecting relevant health care data and categorizing the risk of infections as Low, medium and high using random forest classifier ML algorithm. Fig. 3 shows the various elements of mobile application involving the central data base, interfaces for various stakeholders like health officer, filed workers along with predicted ISP score. The mobile application is interfaced with a versatile database to enable the government administrative, health officer to select individuals for further quarantine and infection elimination protocol execution. The details of the mobile application with the functional modules are given below

**Figure 3.**
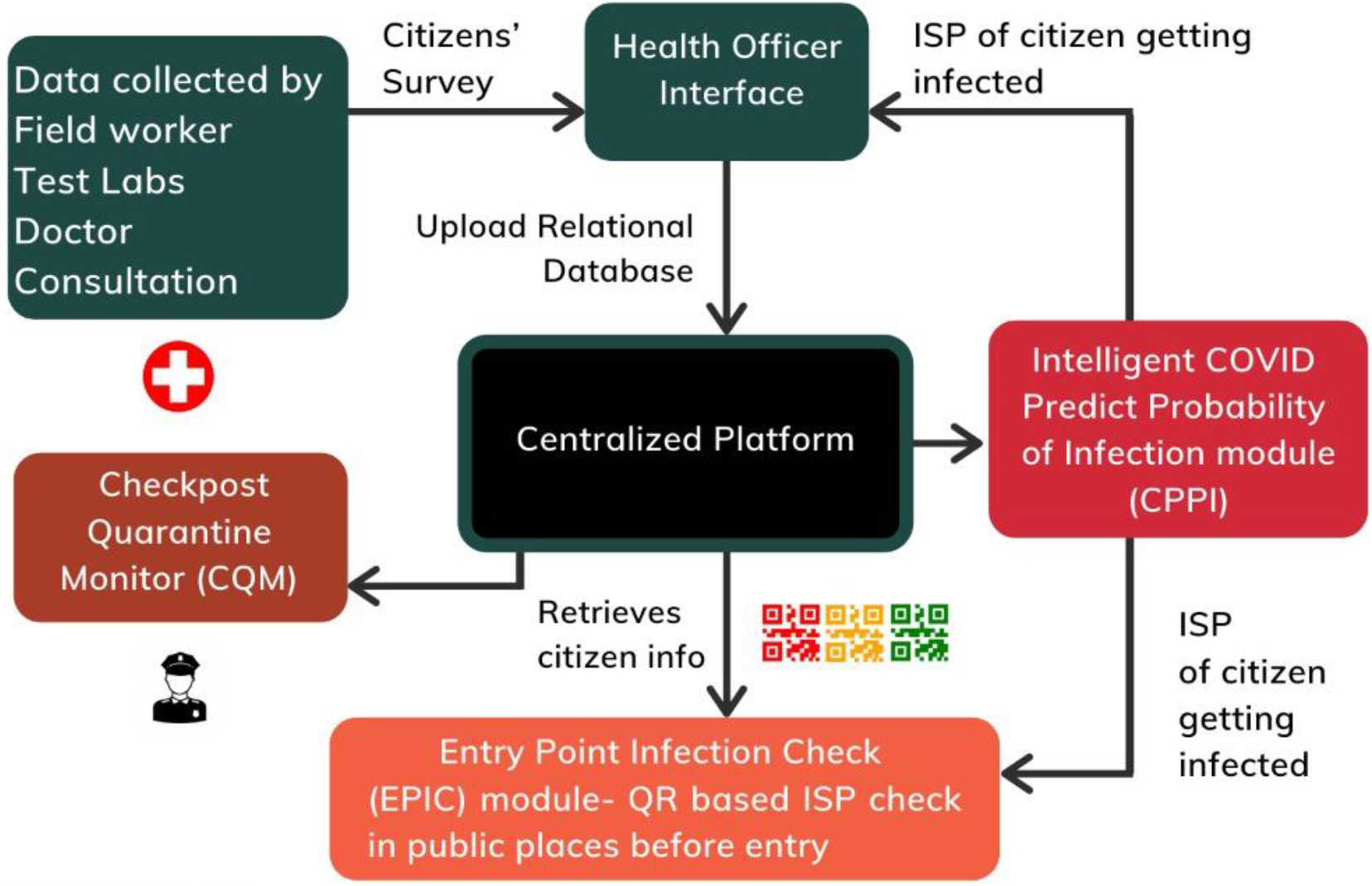
Modules of mobile application platform and different interfaces

1. A centralized data collection system with a graphical user interface in every containment zone, to consolidate and upload data collected from field workers, Doctors and Non-COVID lab tests in a single standardized format.
2. A live geo location tracking that integrates easily with existing open maps and APIs to deliver low power intensive tracking of smartphones using GPS geocoding and decoding techniques.
3. Entry Point Infection Check (EPIC) allows real-time risk assessment through Infection Susceptibility Probability (ISP) score by the proposed machine learning model which produces the QR codes. This can be verified at the public entry points by the concerned Health Officers (HO).

Based on the risk factor score and geo location tracking system in the mobile application, proposed mobile application software tool can be useful contact tracing, tracking of individuals in public places. Also, it can be used for entry point checking tool for screening the individuals based on the risk score of the individuals. The source code for the proposed android mobile application is available in Github digital repository.

### Performance metrics of machine learning techniques

In order to analyze the classification performance of the different machine learning techniques accuracy, Precision, Sensitivity and F-scores are calculated using number of True-Positives, True-Negatives, False-Positives and False-Negatives of the classification.

#### Accuracy

The most common metric for performance assessment measured as a ratio of the number of correctly predicted data to the total number of data provided for testing as calculated by Eq.12.

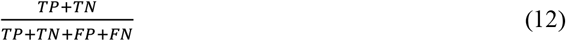

#### Precision

**It is a** quantitative measure to describe the number of correct instances of prediction as compared to the total number of instances provided to the model during testing. This is calculated by eq. 13.

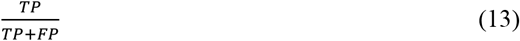

#### Sensitivity

It is measure of ability of the machine learning model to predict the positive labels to all the given labels that should have been predicted positive and it is calculated by eq. 14.

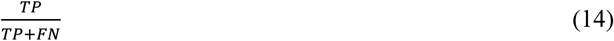

#### F- Score

This is measured by weighted average between precision and recall as calculated by eq15.

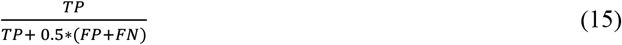

**Here TP= True Positive; TN= True Negative; FP= False Positive; FN= False Negative**

## Results

In the present work, proposed approach is implemented in Python computing and programming environment using the major computing libraries and mathematical functions such as Numpy, Pandas, Scikit learn in Jupyter Notebook environment involving various machine learning algorithms such as random forest, logistic regression, support vector regression, linear regression, and neural networks. An open health care dataset with the infection susceptibility probability score available in the online repository Kaggle is used for training, validation and testing purposes [Srijan Singh, 2020]. The sample health care survey data set is shown in Fig.4. It can be seen that the data set contains categorical data and numerical data.

**Fig.4.**
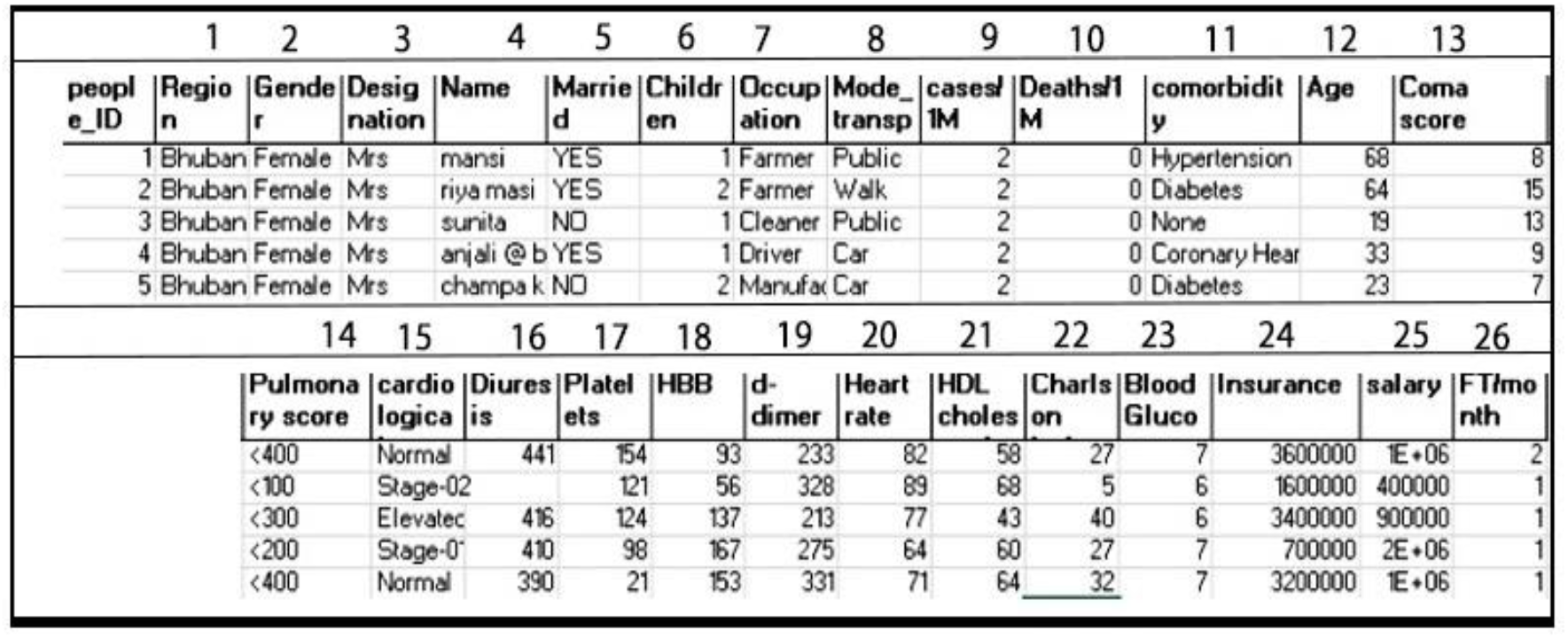
Sample health care data set (Srijan Singh 2020)

The hardware specifications of training, testing and inference include a processor of Intel Xeon CPU at 2.20GHz and a memory of 0.88GB. The given health care data set is preprocessed for overcoming the errors due to the missing data. It was found that random forest model was best among the machine learning models for the supervised classification of target classes of infection probability and the inference timings for 10000 data was found to be 2.85704 seconds.

### Influencing factors of Infection Susceptibility Probability (ISP) score of COVID-19

As the health care data set contains many fields involving epidemiological characteristics, and underlying comorbidities of the individuals, a heat map with a color and magnitude variation is developed to quickly check correlations and visualizing the correlation matrix as shown in Fig. 5.

**Fig 5.**
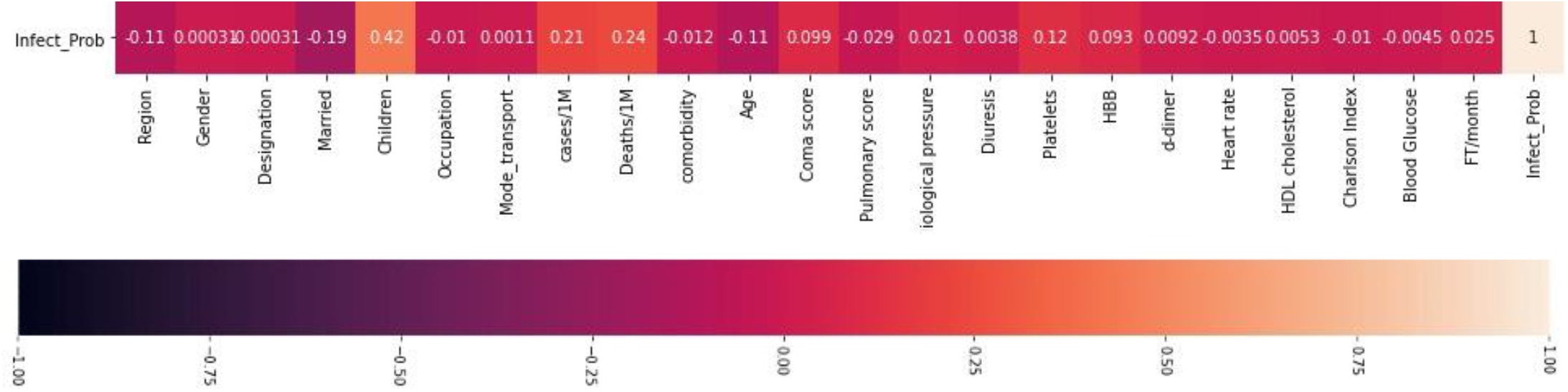
Heat Map of health care data.

This heat map shows the correlation coefficient which varies from +1 to −1 for providing an understanding of the vital and detrimental correlated factors of health care data on the infection probability. From the correlation matrix, four main factors influencing infection probability (ISP) are found to be “No of children”(0.42), “Cases per Million”(0.21), “Deaths per Million”(0.24) and “Platelet Count “(0.12). These results indicate demographic fields are critical in influencing the infection susceptibility score of the individual. It can be seen that the gender and married fields also have an influence on the results which can be quantitatively analyzed from the correlation matrix. Unnecessary fields such as Name, Insurance, Salary, People_ID are found to be non-contributing to our analysis and prediction, thus they are removed during data preparation. We can reduce the correlation matrix of the dataset in which 4 features have negative correlation with the output label.

### Classification performance of RF classifier

In the random forest model, the decision trees developed over 100-200 nodes indicating the complex relationships mapped between the dataset and the target classes such as low, medium, high Infection Susceptibility Probability (ISP) score of COVID-19 infection. A confusion matrix is calculated for understanding the performance of classification of the based on the number of True-Positives, True-Negatives, False-Positives and False-Negatives. These values further used to calculate the accuracy, Precision, Sensitivity and F-scores. Inferring from the confusion matrix as given in Fig.6, proposed model classifies the different groups of people based on their susceptibility with high levels of precision. Here 1 denotes all (100%) the data points belonging to Low and High susceptibility were classified correctly and 0.96 denotes that 96% of the data points were correctly classified under medium susceptibility along with 0.36 denoting 3.6% being misclassified under low susceptibility.

**Fig 6.**
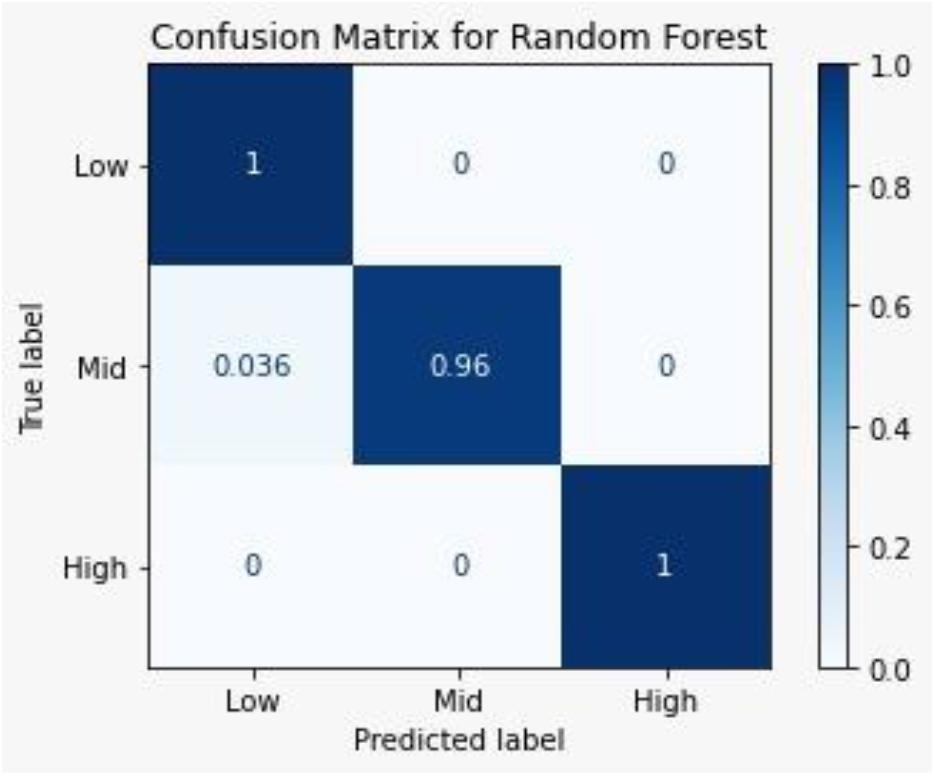
Confusion Matrix for random forest model for the health care data

The performance metrics such as accuracy, Precision, Sensitivity and F-scores are calculated for different machine learning models and it is compared with the RF model as shown in Fig.7. It is found that the random forest approach has an overall classification accuracy of 99.7% for the validation data set of the health care data set. The proposed random forest classifier gives a precision of 99.8%, sensitivity of 98.8% and F-score of 99.29%.

**Fig.7.**
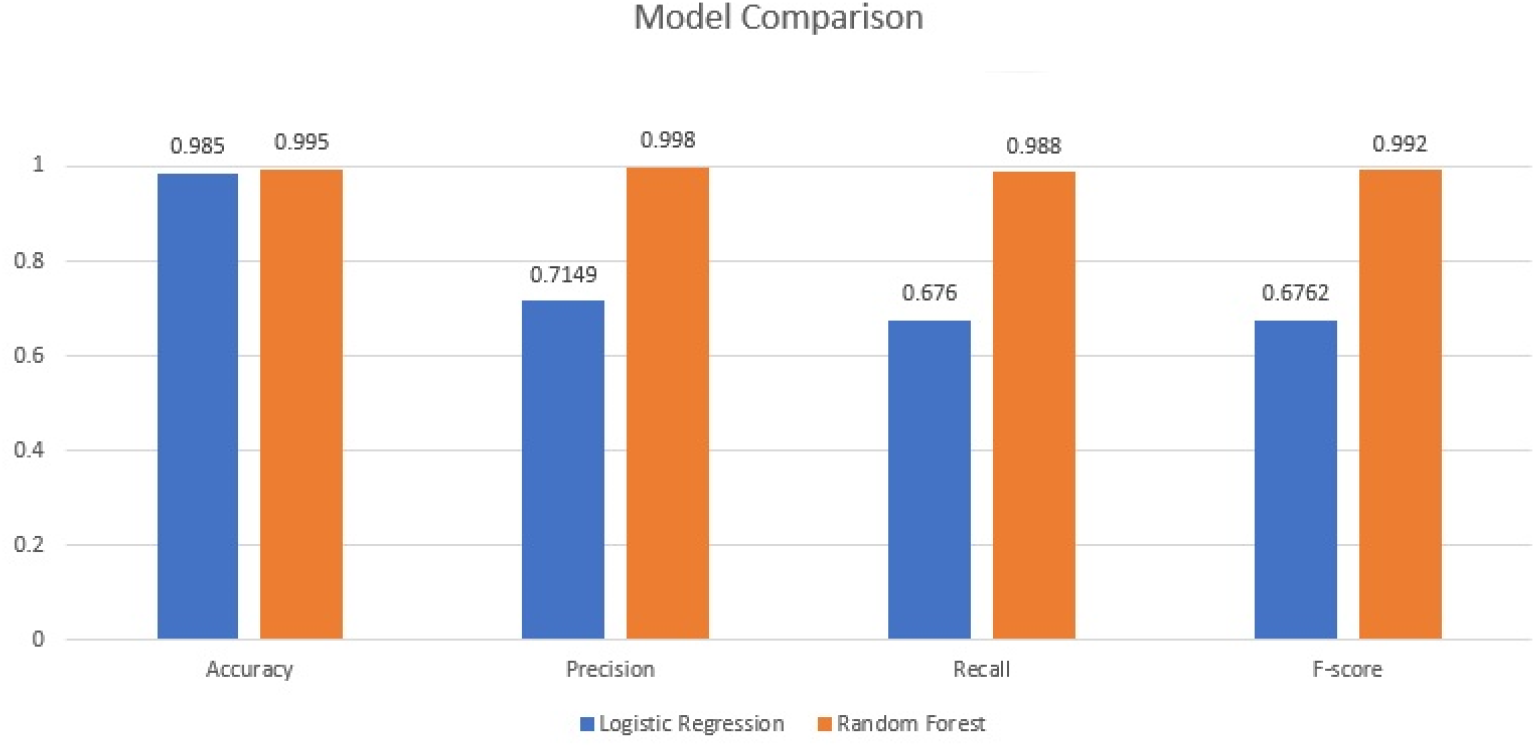
Comparison of performance measure of random forest classifier

The higher sensitivity of random forest model shows that a high proportion of actual positives are classified correctly. Also the higher specificity highlights that a good percentage of the ‘safe’ populations are identified as not susceptible to the infection. The precision indicates that there may be some cases of individuals who may not have a high susceptibility, but they may be declared as a considerably risky individual.

## Discussions

In order to quantitatively analyze the proposed RF model, train and test accuracies over different sizes of samples, scalability of the model, Performance of the model are presented and the results are analyzed

### Effect of size of the data set

In order to study the aspects of over fitting of the proposed random forest model, it is applied to different number of training and testing data sets. This provides an indication of the over fitting of the model and the ability of generalization of classification for the given data set. Fig.8 shows convergence of test and train accuracies and it highlights that the model is not over fitting.

**Fig 8.**
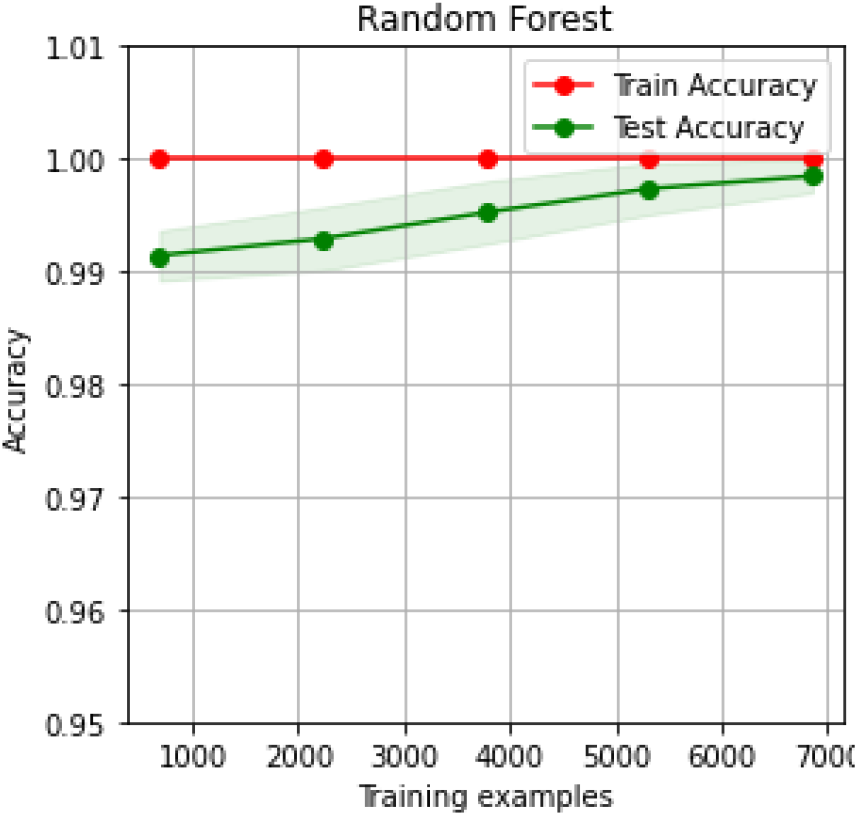
Graph of accuracy of the Random Forest model versus the number of training examples.

The model can produce highly accurate predictions even with a lower number of training examples as the accuracy plot starts at over 99% even for a smaller batch of dataset. This shows the agility of the model to adapt to different dataset sizes without over fitting.

### Scalability Curves

Further, a study has been carried out to understand the scalability of the proposed random forest model, the effect of number of training examples of health care data set on the accuracy of classification and training time. The results are shown in Fig.8 and Fig.9

**Fig 9.**
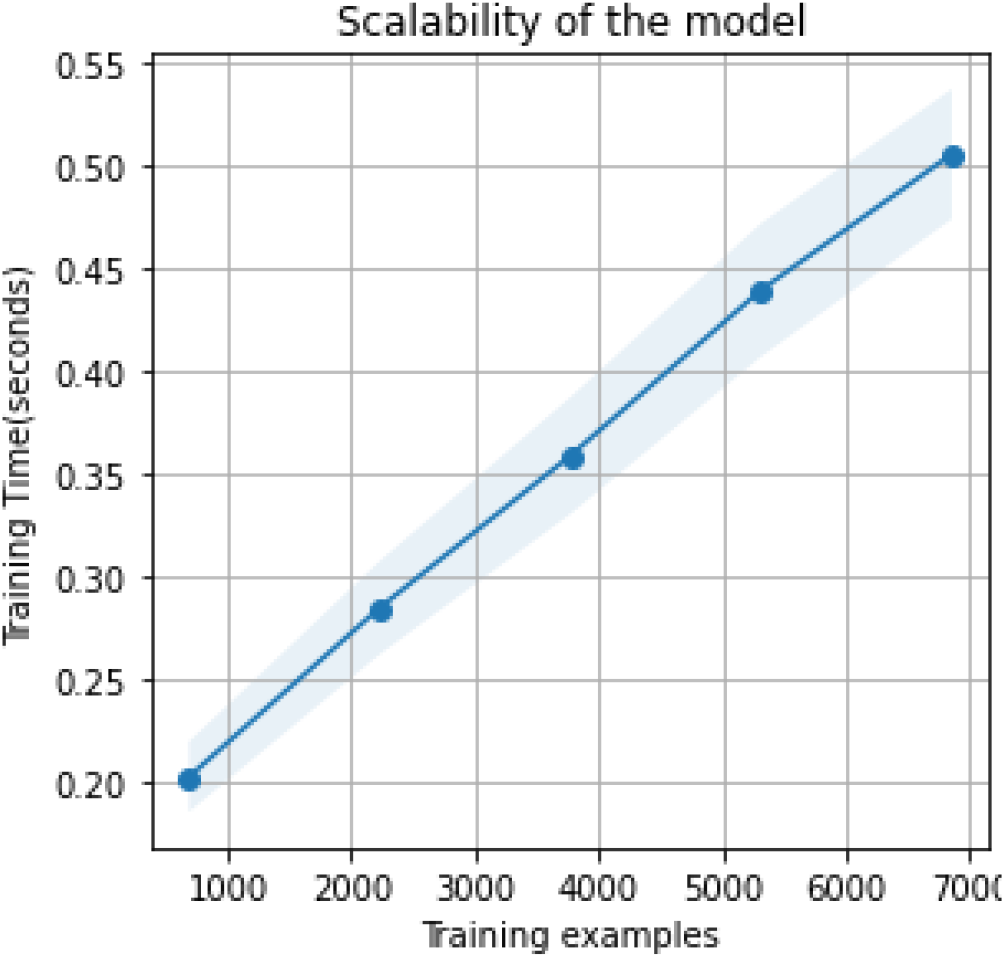
Graph of scalability of the Random Forest model versus the number of training examples

It shows a linear trend for the training time when the size of the dataset is changed, and it indicates the robustness of the model with a higher number of data points.

### Training time

Fig.10 depicts the effect of different numbers of training examples on the training time and it shows the linear trend in accuracy of classification when the size of the dataset is changed.

**Fig 10.**
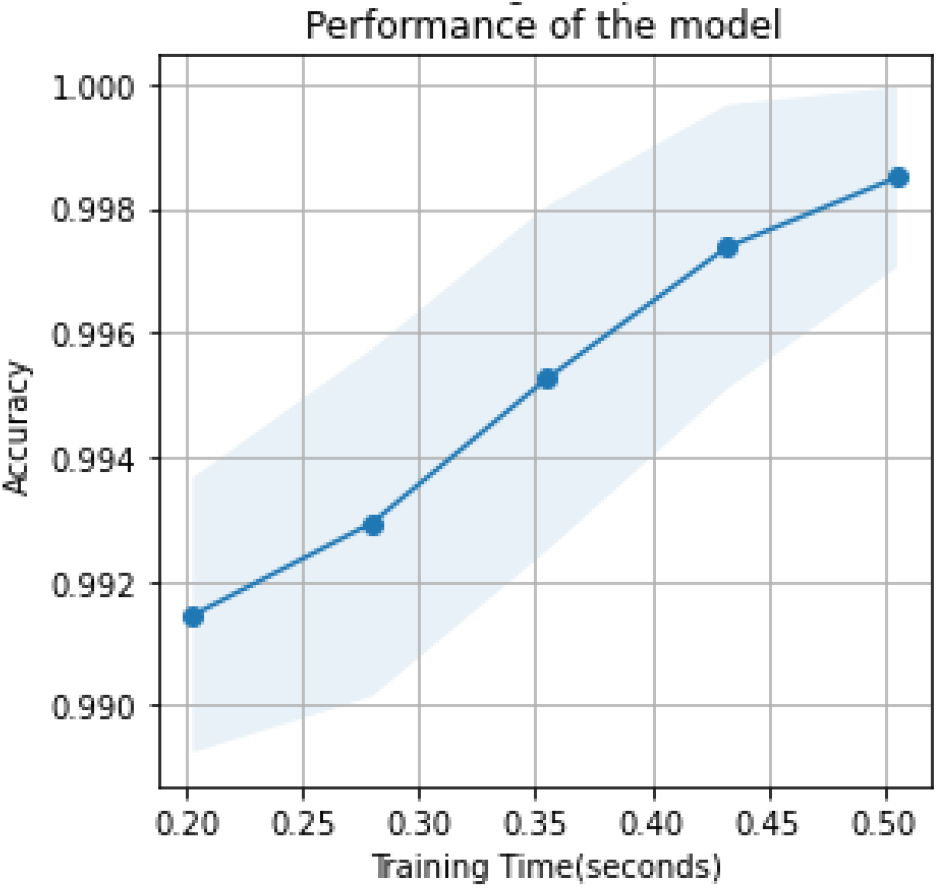
Effect of training time on accuracy of prediction.

It can be seen that there is a smooth increase in accuracy which highlights the robustness of the model with a higher number of training examples. These results indicate that the proposed random forest model performs well with high accuracy despite smaller dataset sizes and requires shorter training time despite large dataset sizes.

### Regression performance of support vector regression

Support vector regression is applied to the health care data set for predicting the infection susceptibility probability score of COVIG-19. From Fig.11(a), it can be seen that there is a lesser deviation between the predicted values and actual values using the RBF kernel function in support vector regression.

**Fig 11.**
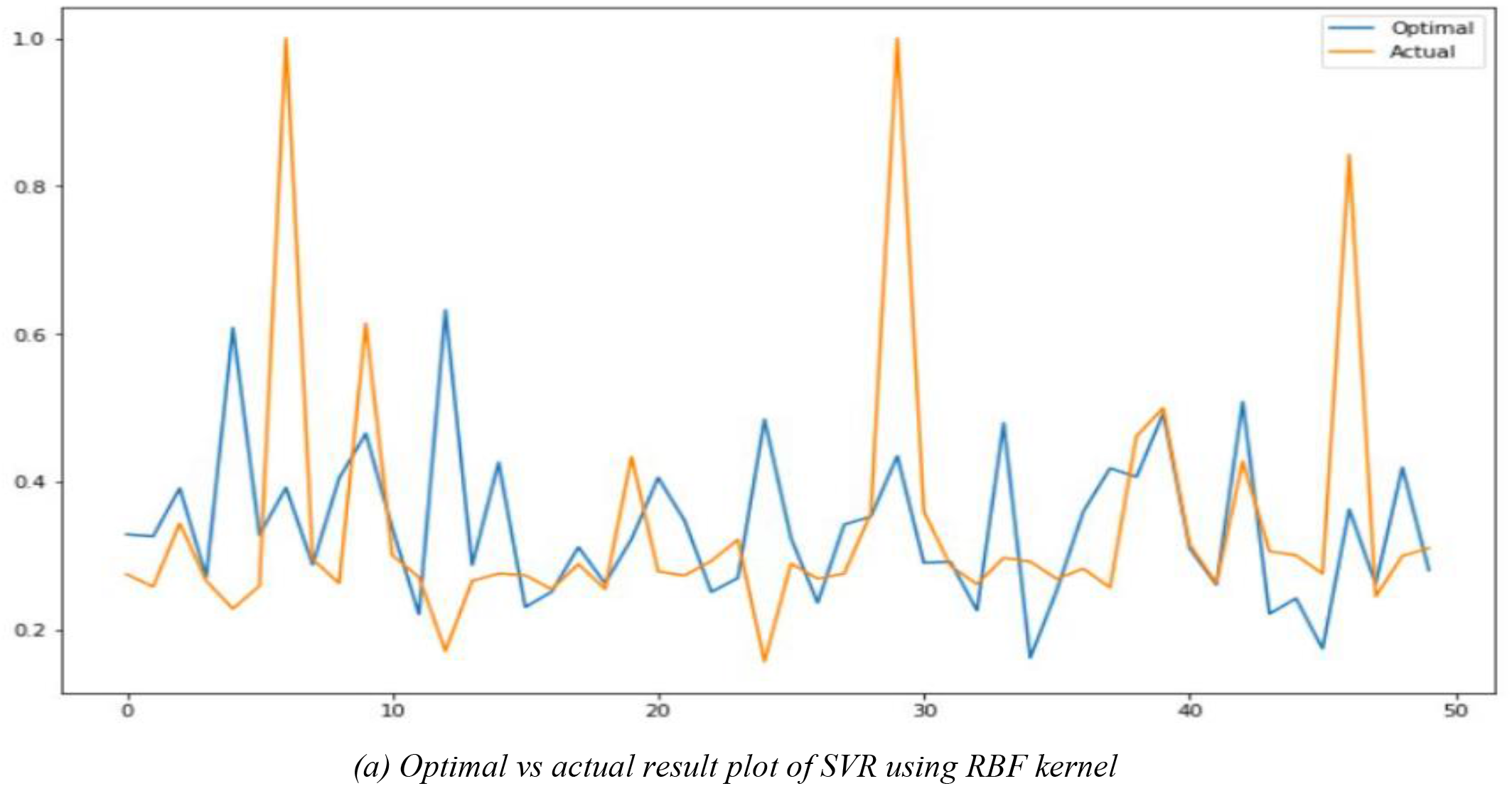
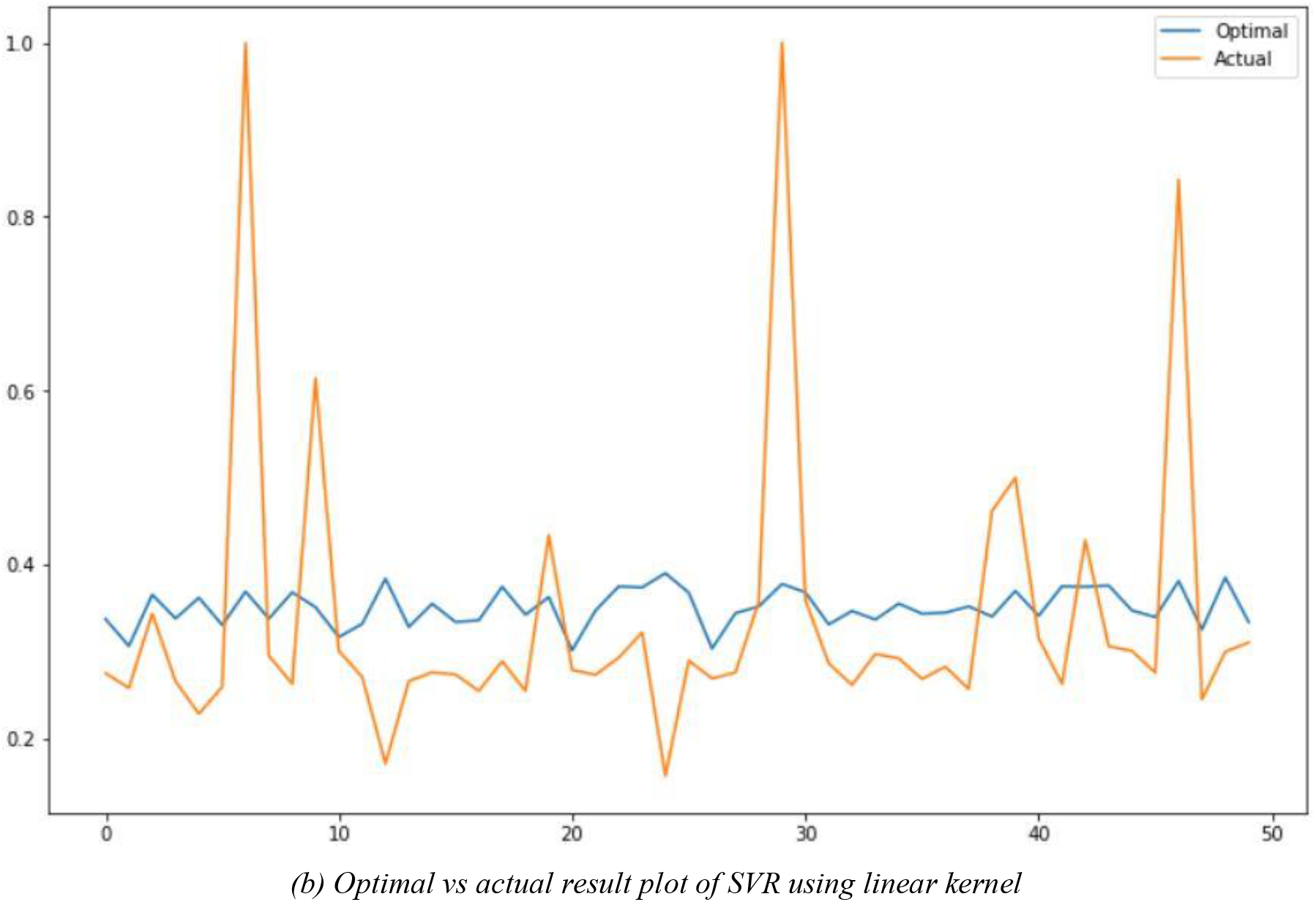
Comparison of Kernel functions used in Support vector regression

Fig.11(b), shows the function fit by the linear kernel of SVR displaying more deviation from the actual values of Infection Susceptibility Probability (ISPs). These results highlights the improved prediction of support vector regression using RBF kernel as compared to the linear kernel

## Conclusions

This paper presented a machine learning based prognostic model for the categorical classification of COVID-19 infection susceptibility as low, medium and high based on the health care data of an individual. A medical data set available in the online repository is used for training, testing and validation of the proposed approach. A mobile application with user interface is developed for collecting relevant health care data and categorizing the risk of infections as Low, medium and high using random forest classifier ML algorithm. Correlation analysis of health care data is carried out to study the influencing factors on influencing the Infection Susceptibility Probability (ISP) score **using the** Heat map and correlation. Demographic fields are found to be critical in influencing the COVID-19 infection such as “No of children”(0.42), “Cases per Million”(0.21), “Deaths per Million”(0.24) and “Platelet Count “(0.12). Based on the comparative study of performance measures for the classification of infection score, it is found that the random forest classifier has the better overall classification accuracy of 99.7% precision of 99.8%, sensitivity of 98.8% and F-score of 99.29% as compared to the other machine learning algorithms such as logistic regression, support vector regression, neural network. Studies on proposed random forest model for different number of data sets highlights robustness and scalability of the proposed approach and it highlights the high accuracy despite smaller dataset sizes and a shorter training time despite large dataset sizes. It is found that support vector regression using RBF kernel function is found to be superior to the linear kernel function in prediction of infection susceptibility of individual for COVID-19.

These results highlighted the application of machine learning approaches for analyzing the health care data in understanding the infection severity of individual for COVID-19. From the larger public health care perspective, proposed approach will be helpful in identification of individuals who are highly susceptible for the COVID-19 infection in a containment zone which can give a decisive role to physicians and government officials for planning the more aggressive treatment and a better chance of survival. Also the early detection can also help hospitals prioritize intensive-care resources.

## Data Availability

The datasets presented in this study can be found online in open source repositories.
Code Availability For the reproducible code, please check out the GitHub repository
at: https://github.com/srivatsanrr/autonom_covid
Following open source repositories are used for implementation of our work: Keras: https://keras.io;
Sklearn: https://scikit-learn.org/stable/. statsmodels: https://www.statsmodels.org/stable/index.html

## Acknowledgements

The authors thank the management of Vellore Institute of Technology, Vellore for providing the necessary facilities to carry out this research work.

## Declarations

### Conflicts of Interest

The authors declare that they do not have any conflicts of interests.

### Funding

There was no funding done for this research work

### Availability of Data and Material

The datasets presented in this study can be found online in open source repositories.

### Code Availability

For the reproducible code, please check out the GitHub Mobile application repository: https://github.com/srivatsanrr/autonom_covid Following open source repositories are used for implementation of our work: Pytorch: https://pytorch.org; Keras: https://keras.io; Sklearn: https://scikit-learn.org/stable/. statsmodels: https://www.statsmodels.org/stable/index.html.

## Notes

### Competing Interest Statement

The authors have declared no competing interest.

### Author Declarations

The research involves no human subjects.

### Summary of Updates

Abstract updated.

